# Risk factors affecting COVID-19 case fatality rate: A quantitative analysis of top 50 affected countries

**DOI:** 10.1101/2020.05.20.20108449

**Authors:** Hui Poh Goh, Wafiah Ilyani Mahari, Norhadyrah Izazie Ahad, Li Ling Chaw, Nurolaini Kifli, Bey Hing Goh, Siang Fei Yeoh, Long Chiau Ming

## Abstract

**Background:** Latest clinical data on treatment on coronavirus disease 2019 (COVID-19) indicated that older patients and those with underlying history of smoking, hypertension or diabetes mellitus might have poorer prognosis of recovery from COVID-19. We aimed to examine the relationship of various prevailing population-based risk factors in comparison with mortality rate and case fatality rate (CFR) of COVID-19.

**Methods:** Demography and epidemiology data which have been identified as verified or postulated risk factors for mortality of adult inpatients with COVID-19 were used. The number of confirmed cases and the number of deaths until April 16, 2020 for all affected countries were extracted from Johns Hopkins University COVID-19 websites. Datasets for indicators that are fitting with the factors of COVID-19 mortality were extracted from the World Bank database. Out of about 185 affected countries, only top 50 countries were selected to be analyzed in this study. The following seven variables were included in the analysis, based on data availability and completeness: 1) proportion of people aged 65 above, 2) proportion of male in the population, 3) diabetes prevalence, 4) smoking prevalence, 5) current health expenditure, 6) number of hospital beds and 7) number of nurses and midwives. Quantitative analysis was carried out to determine the correlation between CFR and the aforementioned risk factors.

**Results:** United States shows about 0.20% of confirmed cases in its country and it has about 4.85% of CFR. Luxembourg shows the highest percentage of confirmed cases of 0.55% but a low 2.05% of CFR, showing that a high percentage of confirmed cases does not necessarily lead to high CFR. There is a significant correlation between CFR, people aged 65 and above (*p* = 0.35) and diabetes prevalence (*p* = 0.01). However, in our study, there is no significant correlation between CFR of COVID-19, male gender (*p* = 0.26) and smoking prevalence (*p* = 0.60).

**Conclusion:** Older people above 65 years old and diabetic patients are significant risk factors for COVID-19. Nevertheless, gender differences and smoking prevalence failed to prove a significant relationship with COVID-19 mortality rate and CFR.

## Introduction

On February 11, 2020, WHO renamed the highly contagious respiratory disease, caused by severe acute respiratory syndrome coronavirus 2 (SARS-CoV-2), as COVID-19 (1). On March 11, 2020, due to the alarming levels of spread and severity of COVID-19 worldwide and by the alarming levels of inaction, WHO has characterized the COVID-19 situation as a pandemic (2). As the COVID-19 outbreak continues to evolve, researchers are learning more about SARS-CoV-2 every day, including the fact that it can be transmitted from symptomatic, pre-symptomatic and asymptomatic people infected with COVID-19 (3). Studies have shown that COVID-19 is primarily transmitted from symptomatic people who are in close contact through respiratory droplets, by direct contact with infected persons or by contact with contaminated objects and surfaces (1). Fever, tiredness and dry cough are the most common symptoms in symptomatic COVID-19 patients (4).

Even though there is limited information regarding risk factors for this severe disease, there are evidences showing a few clear risk factors, including age and underlying medical conditions, might be at higher risk for severe illness from COVID-19 (5,6). Other risk factors that have been discussed included gender, Bacilllus Calmette-Guerin (BCG) vaccination, smoking and malaria prevalence (7,8). Older people have a higher risk due to the decreasing function of the age-dependent lymphocytes resulting in their increased susceptibility to COVID-19 disease (9). In addition, a study shows that there is a higher percentage of death among patients aged over 65 years (62%) than patients aged below 65 years old (37%) (10). Furthermore, male gender is commonly observed in COVID-19 patients (73%) according to a retrospective study done on 113 deceased patients (11).

Another risk factor for COVID-19 mortality is in patients with existing comorbidities. A study by Guan et al. shows that COVID-19 are more commonly seen in patients with hypertension, diabetes, cardiovascular disease and a history of smoking (12). Not only were these patients susceptible to the disease, they also had a higher chance of obtaining poor health outcomes after Immediate Care Unit (ICU) admission and may lead to death (10). Moreover, a study on the correlation between COVID-19 mortality and BCG vaccination suggested that early BCG vaccination could help to decrease the mortality rate (7). Other than that, malaria prevalence is also another risk factor of COVID-19 mortality. According to the research conducted by Spencer, there is a higher number of COVID-19 cases reported in countries with low malaria prevalence than countries that had higher malaria prevalence (13). Apart from addressing risk factors, there are also parameters that may affect the COVID-19 mortality rate such as shortage of staff, lack of medical supply or equipment, insufficient hospital beds and the country’s health expenditure.

As of end of April 2020, SARS-CoV-2 virus has resulted in more than 3.1 million infections and over 217,000 deaths globally (1). As COVID-19 has become a global pandemic issue, implementation of suitable interventions will be needed for the public, healthcare professionals and patients and to ensure all sectors to work together cohesively and efficiently. Even though COVID-19 origins from coronavirus, the SARS-CoV-2 has very different severity and contagion characteristics and much still needs to be learned about it. Thus, it is imperative to evaluate the relationship of postulated or verified risk factors with COVID-19 mortality. It is absolute crucial to evaluate the risk factors of mortality among patients infected with COVID-19 at population level. By validating the relationship, patients with COVID-19 can be treated more aggressively than those without the risk factor. The findings of the current study provide a clinical picture of COVID-19 case fatality rate in top 50 affected countries. A predictive model of COVID-19 population infection rate and case fatality rate that will increase clinicians’ attention for this highly fatal and morbid disease was developed. These findings consolidate the evidence of crucial risk factors that front liners that need to prioritise to decrease the COVID-19 mortality globally.

## Methods

### Data extraction

Demography and epidemiology data which have been identified as verified or postulated risk factors for mortality of adult inpatients with COVID-19 were used. The data were obtained from those published in the World Bank and Johns Hopkins University COVID-19 websites. The number of confirmed cases and the number of deaths for all affected countries were extracted from the latter (14). Datasets for indicators that are fitting with the factors of COVID-19 mortality were extracted from the World Bank database (15). Data extracted for this study is up until April 16, 2020. All data acquired were exported in excel format and arranged according to country rankings with the top having the highest number of confirmed cases as of April 16, 2020 and the bottom having the least. Out of about 185 affected countries, only top 50 countries were selected to be analyzed in this study.

The following seven variables were included in the analysis, based on data availability and completeness: 1) proportion of people aged 65 above, 2) proportion of male in the population, 3) diabetes prevalence, 4) smoking prevalence, 5) current health expenditure, 6) number of hospital beds and 7) number of nurses and midwives.

### Data analysis

For each country, the percentage of confirmed COVID-19 case per country was calculated by dividing the number of confirmed COVID-19 cases by the total population for each country. Also, CFR was calculated by dividing the number of deaths related to COVID-19 by the confirmed COVID-19 cases. Bar graphs are plotted to illustrate both measures.

Regression analysis was conducted to determine the risk factors of CFR for COVID-19. For this analysis, few variables (CFR and number of hospital beds) were standardized due to differences in scale and very large range. Standardization was done by subtracting each value by the mean and then dividing it with the standard deviation. Also, some variables (diabetes prevalence, current health expenditure, and number of nurses and midwives) were divided into four equal categories (i.e. in quartiles). All analyses were conducted using Microsoft Excel and R (ver. 3.6.0). A *p*-value < 0.05 was considered as statistically significant.

## Results

Information of 2,017,444 confirmed COVID-19 cases and 137,166 deaths from each of the 50 top countries (Supplementary Information) were extracted. This constitutes about 93.3% and 92.8% of the global confirmed cases and deaths on the data collection date (April 16, 2020).

### Case Fatality Rate (CFR) and mortality rate of COVID-19 in the top 50 countries

From data extracted up until April 16, 2020, the United States (US) reported to have the highest number of total confirmed cases and the highest total number of deaths of 639,733 cases and 31,002 cases respectively. Despite that, the US shows about 0.20% (Table 1) of confirmed cases in its country and it has about 4.85% of CFR (Table 1), indicating a moderate death rate of COVID-19 in comparison to other countries. Luxembourg, ranked 47^th^ in the list of top 50 COVID-19 countries, shows the highest percentage of confirmed cases of 0.55% (Table 1) but a low 2.05% of CFR (Table 1), showing that a high percentage of confirmed cases does not necessarily lead to a high CFR. This is due to variations in number, transmission rate and severity of the disease regardless of the rankings. Hence, it is important to evaluate the possible factors that can affect the increase of COVID-19 mortality rate globally.

**Table 1:** Percentage of confirmed CFR and COVID-19 cases (In sequence of the highest to lowest CFR%)

| No. | Countries | Confirmed COVID-19<br>cases (%) | Case Fatality Rate (%) |
| --- | --- | --- | --- |
| 1 | Belgium | 0.3 | 13.95 |
| 2 | United Kingdom | 0.15 | 13.82 |
| 3 | Italy | 0.27 | 13.11 |
| 4 | France | 0.2 | 12.77 |
| 5 | Netherlands | 0.17 | 11.32 |
| 6 | Sweden | 0.12 | 10.63 |
| 7 | Spain | 0.39 | 10.46 |
| 8 | Indonesia | 0.002 | 8.99 |
| 9 | Mexico | 0.005 | 7.68 |
| 10 | Philippines | 0.01 | 6.4 |
| 11 | Iran | 0.1 | 6.24 |
| 12 | Brazil | 0.01 | 6.07 |
| 13 | Dominican Republic | 0.03 | 5.23 |
| 14 | Romania | 0.04 | 5.09 |
| 15 | Ecuador | 0.05 | 4.94 |
| 16 | United States | 0.2 | 4.85 |
| 17 | Switzerland | 0.31 | 4.8 |
| 18 | Denmark | 0.12 | 4.54 |
| 19 | Colombia | 0.01 | 4.22 |
| 20 | China | 0.01 | 4.01 |
| 21 | Poland | 0.02 | 3.76 |
| 22 | Canada | 0.08 | 3.56 |
| 23 | Ireland | 0.26 | 3.54 |
| 24 | India | 0.001 | 3.4 |
| 25 | Portugal | 0.18 | 3.33 |
| 26 | Germany | 0.1 | 2.86 |
| 27 | Ukraine | 0.01 | 2.79 |
| 28 | Panama | 0.09 | 2.75 |
| 29 | Austria | 0.16 | 2.73 |
| 30 | Czech Republic | 0.06 | 2.63 |
| 31 | Finland | 0.06 | 2.23 |
| 32 | Norway | 0.13 | 2.21 |
| 33 | Peru | 0.04 | 2.21 |
| 34 | Turkey | 0.08 | 2.19 |
| 35 | Korea, Rep | 0.02 | 2.16 |
| 36 | Japan | 0.01 | 2.06 |
| 37 | Luxembourg | 0.56 | 2.05 |
| 38 | Serbia | 0.07 | 2.03 |
| 39 | Pakistan | 0.003 | 1.85 |
| 40 | Thailand | 0.004 | 1.72 |
| 41 | Malaysia | 0.02 | 1.62 |
| 42 | Saudi Arabia | 0.02 | 1.3 |
| 43 | Chile | 0.04 | 1.14 |
| 44 | Israel | 0.14 | 1.11 |
| 45 | Australia | 0.03 | 0.97 |
| 46 | Belarus | 0.04 | 0.95 |
| 47 | Russian Federation | 0.02 | 0.83 |
| 48 | United Arab Emirates | 0.06 | 0.62 |
| 49 | Singapore | 0.07 | 0.27 |
| 50 | Qatar | 0.15 | 0.17 |

### Relationship between the different risk factors and COVID-19 CFR

The proportion of people aged 65 and above has a significant association with CFR (*p* = 0.04, Table 2). The β coefficient of 4.70 tells us that for every 1-unit increase in the proportion of people aged 65 years, the CFR increases by 4.7 units. This relationship is illustrated in Figure 1, where CFR sharply increases when the proportion of people aged 65 years is 0.15 and above. Also, there is a certain association between diabetes prevalence and CFR. When compared to countries with low prevalence (31. – 5.5%), those with highest prevalence (9.67-19.9%) have lower CFR by 0.97 units (*p* = 0.01, Table 2). However, no significant association was observed when including both variables (proportion of people aged 65 and above and diabetes prevalence) into the multiple regression analysis.

**Table 2:**
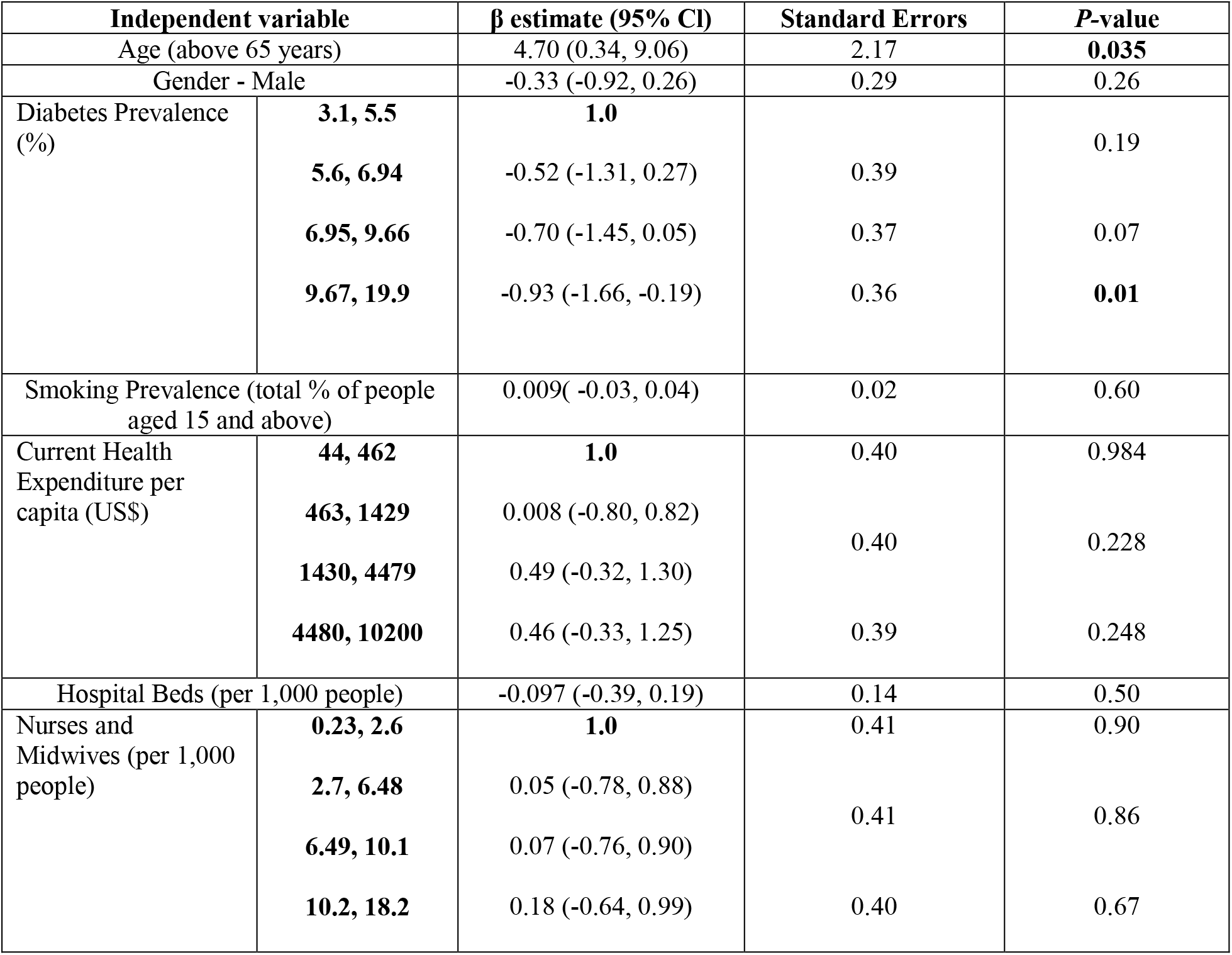
Correlation between different risk factors and COVID-19 CFR

**Figure 1:**
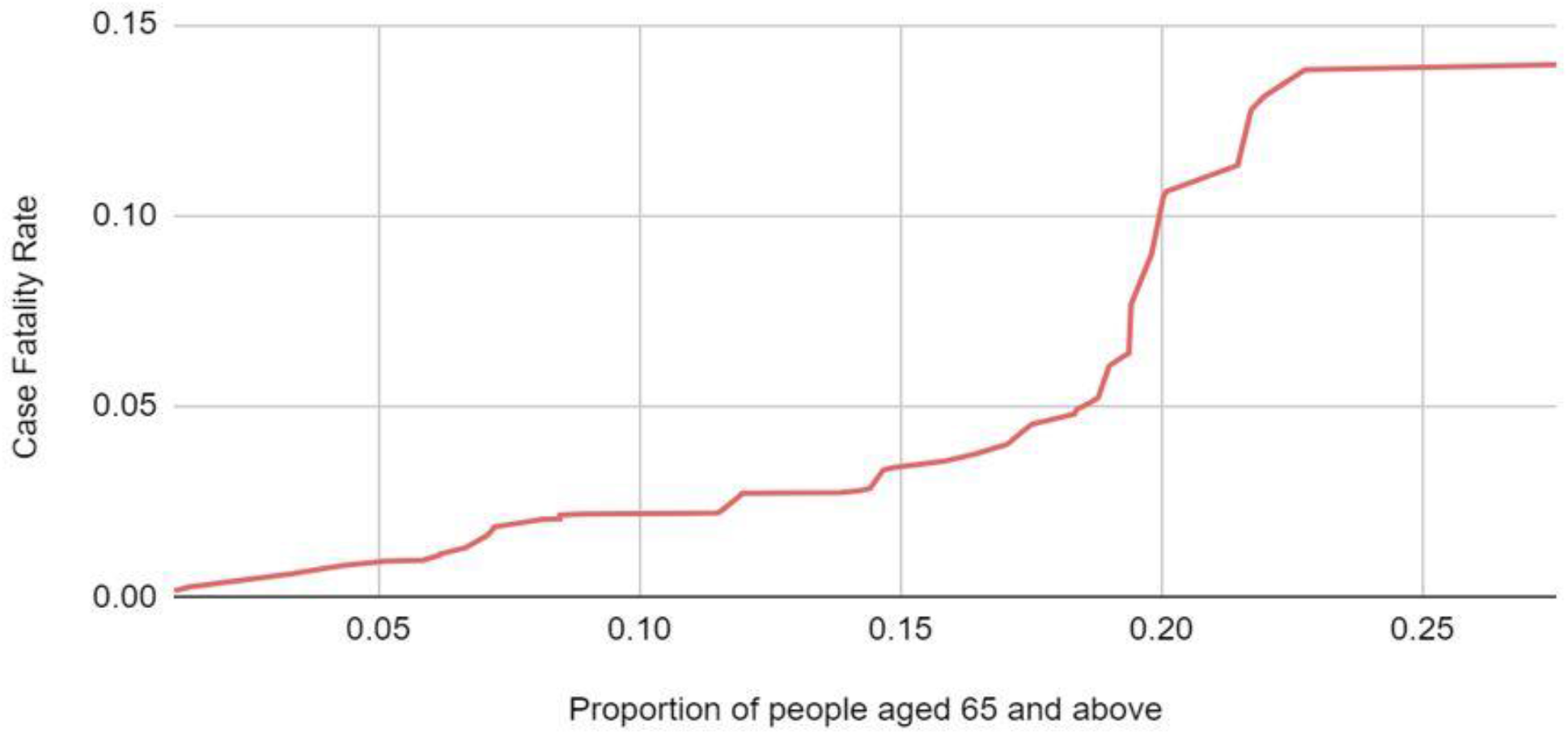
Correlation between case fatality rate and proportion of people aged 65 and above

## Discussion

There are still a lot of unknown regarding the disease COVID-19. There is a steep learning curve about the virus, and this could take a couple of years to work out. However, we are not completely in the dark when it comes to risk factors.

Studies have shown that age is a clear risk factor for severe COVID-19 disease and thus, resulting in death. This has been confirmed by our study where the proportion of people aged 65 and above has shown a significant correlation with CFR. This indicates that countries with a higher proportion of people aged 65 and above may result in higher COVID-19 mortality rate. Bhatraju et al. (2020) has shown that in Seattle, the US reported more than 60% of COVID-19 deaths in patients aged 65 years and above than those who are younger than 65 years old (10). Verity et al. (2020) has shown that the case fatality rate for those under age 60 was 1.4% while the fatality rate increases drastically to 4.5% for those people aged over 60 years old (6). This shows that the older the population, the higher the fatality rate. For those 80 years old and over, COVID-19 appears to have a 13.4% fatality rate (6). Furthermore, deceased patients were found to be at an average age of 68 years old while recovered patients to be at an average age of 51 years old (11). These studies show that COVID-19 disproportionately impacts certain groups, and older people is one of the vulnerable groups. There is no one reason to this; it is believed that immune system declines with age. An increase of deficiency in T-cell and B-cell function and overproduction of type 2 cytokines as age increases (9). This may increase the viral replication and extend the duration of pro-inflammatory responses leading to poor health results (9). Older people tend to have more underlying conditions that may also be risk factors for severe COVID-19 (6,12).

Even though there is no significance shown between COVID-19 CFR and the male gender, it is important to note that differences in gender may play a role in severity of COVID-19. There are studies that have shown COVID-19 affecting more males than females (16-18). This could be due to males having more underlying health risk factors than the female population or the fact that males tend to engage in more risky health-relatable behaviours, such as greater rates of smoking and drinking alcohol (19). Genetics and differences in immune response can be explanations to this phenomenon too.

Studies have shown that many of the severe COVID-19 patients also have underlying medical conditions, such as diabetes and cardiovascular diseases (11,20). Our study has confirmed that there is indeed a certain association between diabetes prevalence and CFR. However, it is important to note that according to our study, diabetes prevalence may be an “unreliable” variable as it was shown that countries with high diabetes prevalence have lower COVID-19 CFR than countries with low diabetes prevalence. Further investigation is needed to define the actual association between diabetes prevalence and COVID-19. Although smoking prevalence has shown no significant association with COVID-19, it cannot be assumed that there is no correlation between other co-morbidities and COVID-19 CFR since not all factors were considered in this study, such as hypertension and cardiovascular diseases (18). Patients with existing comorbidities, including hypertension, diabetes, cardiovascular disease and history of smoking, seems to be associated with COVID-19 more severely (12). With reference to a retrospective study of 113 deceased patients from COVID-19, 48% of the patients had chronic hypertension and 14% of them had cardiovascular diseases (11). In addition to that, COVID-19 patients who have hypertension were closely associated with poor health outcomes after hospital admission. This may be due to factors such as vascular aging, reduced renal function and medication interactions (21).It is important to note that everyone is responsible in controlling this COVID-19 pandemic, slowing the rate of transmission and hence successfully flattening the curve. An increase in COVID-19 cases can affect the country’s economy. It is necessary for some countries may need more supplies than others to cater all sick patients, and thus increasing the health expenditure of the country (2). Some of the medical supplies include Personal Protective Equipment (PPE), mechanical ventilators, COVID-19 testing kits and Extracorporeal Membrane Oxygenation (ECMO). Due to surge of demand on health care system, countries with low income and poor healthcare infrastructure suffer the most (12). However, in our study, current health expenditure does not significantly correlate to an arising COVID-19 CFR. One possible explanation to this is the various levels of well-preparedness plans implemented in the country to handle this pandemic situation. These include availability of medicines or medical supplies to treat COVID-19 patients, availability of suitable places to quarantine or self-isolate these patients, number of healthcare professionals and a strategized interventions, such as social distancing, quarantine, isolation actions and proper management, for the patients and public to flatten the curve and to reduce healthcare burden.

Even though there is no significant correlation, this does not indicate health expenditure is not important. Hospital beds is an example of hospital expenditures, and is important in the improvement of hospitals by making adjustments in volume-based to fulfill healthcare demands (2). Improvement in hospital capacity is necessary to accommodate any future pandemic situations (22). Despite the insignificant correlation to COVID-19 CFR, accessibility to adequate hospital beds for COVID-19 patients can potentially affect the CFR as the number of beds required depends on the number of confirmed cases in each country. Apart from hospital beds, other medical supplies such as PPE and mechanical ventilators must be sufficient as they are the key equipment for frontline healthcare workers (23-25). Evidence of high numbers of infections and deaths among healthcare workers due to lack of PPE were reported in Italy (23). In the US, recent estimates have suggested that the estimated ventilators needed is ranging from several hundred thousand to a million, far more than what are currently available (23). It is difficult to estimate the exact number of ventilators needed as it depends on the number, transmission rate and severity of the disease in each country. Although there is no significant relationship between the number of nurses and midwives and CFR, number of healthcare professionals is a crucial factor in the COVID-19 outbreak. The reason to this insignificance can be due inadequate data obtained as the data collected are only numbers of nurses and midwives. It is important to take into account the numbers of other healthcare professionals. Physicians and nurses who had no disease expertise were also recruited to provide care to patients with COVID-19 (25). Recognizing the risk of healthcare workers shortages and ensuring them to have adequate rest and to feel supported are important to maintain individual and team performance in overcoming challenges presented by COVID-19 (24).

There are a number of limitations in this study that need to be acknowledged. Firstly, some factors had to be excluded due to incomplete data such as malaria prevalence and BCG vaccination. Secondly, the years from which the data was collected were not consistent for all indicators. Thirdly, the data collected were not from the same year for one indicator such as the number of hospital beds. Lastly, some required data were unavailable to sufficiently make an overall conclusion for some of the factors, including comorbidities. There were 4 other proposed comorbidities to be analyzed but only two indicators’ datasets were available in World Bank Data, which are diabetes and smoking prevalence. Therefore, more research should be conducted to further understand the relationship between comorbidities and CFR. This would help to identify and to better understand other possible factors that may also affect CFR.

## Conclusion

As COVID-19 is such a new disease, much still needs to be learned about it. Age is a clear risk factor for severe COVID-19 and death. COVID-19 is an illness that disproportionately impacts older people. However, aforementioned risk factors should not be neglected as they may play essential roles in flattening the curve and reducing healthcare burden. Prediction alone is not efficient, but well-planned and suitable interventions should also be carried out. In addition to that, potential risk factors need a lot more research in order to understand the risks for the worst forms of COVID-19 and what we ought to learn to best protect the most vulnerable people. Participation and involvement of every individual, including patients, public and healthcare professionals, is necessary and everyone should work together towards combating COVID-19 disease.

## Data Availability

The authors confirm that the data supporting the findings of this study are available within the article and its supplementary materials.

## Author contribution

LCM conceived the project. HPG, WIM, NIA, LLC, LCM analyzed results and interpreted the data and wrote the manuscript draft. NK, BHG, SFY revised the manuscript. All authors read and approved the final manuscript.

